# DrugOn: A Comprehensive Drug Ontology for Precision Oncology

**DOI:** 10.1101/2024.09.23.24314201

**Authors:** Kevin Kornrumpf, Vera Gnaß, Myrine Holm, Tim Beißbarth, Raphael Koch, Jürgen Dönitz

## Abstract

**Introduction:** Precision oncology and biomedical cancer research increasingly rely on tools to select optimal drugs targeting specific genetic alterations in cancer. A major challenge for bioinformatic tools supporting drug selection is to standardize different annotations (substance, drug name, drug class) to a common level, typically the drug class. While manual classification is time-consuming and potentially biased, existing resources often lack completeness, granularity, or mix up drug classes and drug targets. A structured, automatically built drug ontology such as DrugOn fills these gaps, improving decision support and data-driven research.

**Methods:** DrugOn integrates information from multiple sources to create a comprehensive drug ontology. It includes categories, molecular targets and additional annotations from DrugBank, ATC, MesH, KEGG and ClueIO. The combination of this data enables accurate identification of drug categories for each drug.

**Results:** DrugOn’s effectiveness was demonstrated by classifying 336 drugs from CIViC. It agreed with manually curated database-derived classifications for 268 out of 282 drugs assessed in translational lymphoma research. In 54 cases, classification was not possible due to data gaps. DrugOn provides a REST API and a front-end application for ontology exploration and automated drug queries.

**Conclusion:** DrugOn, a unified drug ontology, is derived from public datasets and refined by precise processing rules to ensure a reliable, updatable resource for drug information, in precision medicine. It uniquely categorizes drug classes and target proteins, and its structured format, complemented by an accessible API, allows for easy integration into data driven pipelines. Initially tailored for lymphoma research, DrugOn’s adaptable nature supports broader cancer research applications and potential data source expansions. DrugOn is accessible at https://mtb.bioinf.med.uni-goettingen.de/drugon-web.

## Introduction

Precision medicine represents a shift in healthcare that enables personalized treatment options based on specific genetic signatures. This approach relies on a comprehensive analysis genetic alterations, resulting in a larger volume of patient-specific data [1]. To effectively use this data, clinicians typically query multiple databases and clinical trial repositories. The gathered information about specific drugs span a wide range of formats and levels of detail, including drug names, brand names, active compounds, target pathways, approval numbers, and accession codes. Handling of drug information is important for the preparation of Molecular Tumor Boards (MTBs) on the one hand, and data-driven wet lab projects on the other [2, 3]. Both rely on the automatic processing of normalized data and the mapping to the common level of a drug class.

To retrieve drug categories, traditional classification methods such as the Anatomical Therapeutic Chemical (ATC) Classification System (https://atcddd.fhi.no/)and Medical Subject Headings (MeSH) provide information in a one-dimensional, hierarchical view. However, the hierarchical levels of these resources are often not related to the classes that are used by clinicians. The Kyoto Encyclopedia of Genes and Genomes (KEGG) [4], ClueIO [5, 6] and comprehensive repositories such as DrugBank [7–12] offer a unique perspective on drug categorization. Drug-Bank, in particular, is also rich in detailed information on most of the known drugs and chemical compounds, including their targets and mechanisms of action.

With the rapidly evolving field of precision medicine the amount of data to process and the number of approved drugs increased in the last ten years [13, 14]. The need for comprehensive and integrative data structures is more relevant than ever. However, the main challenge is to integrate these different sources into a coherent, accessible framework.

Ontologies are the state of the art method to collect and organize a complex resource like drug information. They consist of ontology classes representing concepts of the domain and a collection of named connections to set them in relation. The resulting graph represents the complex connections between the included entities. They represent an ideal data structure, as they create a standardized language and framework for categorizing and linking information. This enables efficient integration and comparison of drug data from various sources, by organizing related information, such as brand names, mechanisms of action and targets.

The need for a unified drug ontology in precision medicine is underscored in data-driven projects, where drugs must be accurately assigned to their respective classes, mechanisms of action, and targets. Current practices often involve manual annotation of this information, a process that is not only work intensive and time-consuming, but also prone to errors and inconsistencies [15]. Moreover, the deterministic nature of this task is often questioned, given the subjective biases that can influence the decision-making process [16–18].

Already existing ontologies such as the Drug Repurposing Ontology (DRON) [19], the Systematized Nomenclature of Medicine (SNOMED) [20], and RXNorm [21] provide extensive data. However, they do not always meet the specific needs of precision medicine, which requires a more refined and meaningful approach, as these databases do often not cover all the relevant levels and connections, limiting their value in other applications.

To fill this gap, we are introducing DrugOn, a comprehensive drug ontology. DrugOn is designed to integrate and normalize the variety of data available from multiple sources to provide a multidimensional hierarchical view of drugs. Unlike static databases, DrugOn is an easily extensible and maintainable framework. With automatic updates it keeps pace with the rapid advances in drug research and knowledge.

DrugOn also enhances usability with an interactive graphical interface. This visual exploration helps identify complex associations and patterns, making DrugOn an helpful tool for researchers, clinicians, and data scientists.

The ontology is built primarily using the Web Ontology Language (OWL) and integrates several data resources. OWL is an open, well known data-format and facilitates a rich and complex ontology structure for effective representation of drug information.

In summary, the development of DrugOn is an important building block in applications supporting precision oncology. By addressing the limitations of existing systems and introducing features tailored to the needs of precision medicine, DrugOn sets a new standard for drug classification, drug repurposing and identifying potential alternative treatment strategies in precision medicine. Its dynamic, integrative and user-friendly design ensures that it will be a valuable resource in personalized medicine.

## Materials and methods

### Data Resources

The data provided by DrugOn is primarily derived from a variety of established data sources, ensuring comprehensive coverage and reliability. These sources include the Anatomical Therapeutic Chemical (ATC) Classification System, the Medical Subject Headings (MeSH) database, the Kyoto Encyclopedia of Genes and Genomes (KEGG), with a particular focus on drug groups and classes, the ClueIO (version 2018-09-07) database, and the DrugBank database (version 5.1.12, 2024-03-14) [4–12]. These platforms provide a wide range of information, from drug classifications to molecular details, contributing to the scope and power of the ontology.

For validation, we conducted a comprehensive analysis of all therapeutic cancer drugs listed in the Clinical Interpretation of Variants in Cancer (CIViC) database [22, 23]. Each drug has been annotated with insights from clinical expertise and a comprehensive literature search, and has been specifically assigned to the appropriate drug category in the clinical context (see Table 1).

**Table 1.** Excerpt of manual drug classification. The table shows a selection of drugs annotated with the categories and with DrugOn’s. The first 15 entries show matching annotations for the manual and DrugOn classification, while the last 5 entries show differences due to incorrect interpretations or missing background information on the part of DrugOn.

| Drug | Drug Category - manual | Drug Category - DrugOn | Match |
| --- | --- | --- | --- |
| Abemaciclib | CDK4/6 inhibitor | cdk inhibitor | ✓ |
| Afatinib | EGFR inhibitor<br>HER2 inhibitor<br>HER4 inhibitor | egfr inhibitor | ✓ |
| Alectinib | ALK inhibitor | alk tyrosine kinase-receptor inhibitor | ✓ |
| Alemtuzumab | CD52 antibody | cd52 antibody | ✓ |
| Avapritinib | KIT inhibitor<br>PDGFR inhibitor | kit inhibitor | ✓ |
| Axitinib | PDGFR inhibitor<br>VEGF inhibitor | pdgfr tyrosine kinase-receptor inhibitor<br>vegfr inhibitor | ✓ |
| AZD3463 | ALK inhibitor | alk tyrosine kinase-receptor inhibitor<br>insulin growth factor-receptor inhibitor | ✓ |
| AZD5363 | AKT inhibitor | akt inhibitor | ✓ |
| BCL2 inhibitors | BCL2 inhibitor | bcl2 inhibitor | ✓ |
| BET inhibitors | BET inhibitor | bet inhibitor | ✓ |
| BRAF inhibitor | RAF inhibitor | braf inhibitor | ✓ |
| BRAF inhibitors | RAF inhibitor | braf inhibitor | ✓ |
| BRAF inhibitors In BRAF Mutant Tumor | RAF inhibitor | braf inhibitor | ✓ |
| Cetuximab | EGFR inhibitor | antineoplastic-egfr inhibitor | ✓ |
| Chemotherapy |  | photochemotherapy | ✗ |
| MEK inhibitors In BRAF Mutant Tumors | MEK inhibitor | braf inhibitor | ✗ |
| corticocorticosteroids In Early Setting | corticosteroid |  | ✗ |
| Pan-RAF inhibitor | RAF inhibitor | pan inhibitor | ✗ |
| Birabresib | BET inhibitor |  | ✗ |

### General selection of drug categories

The drug classification methodology utilized by DrugOn is a complex, multi-step approach that is designed to ensure the accuracy of drug categorization based on a variety of criteria. The complete workflow is illustrated in Figure 1.

**Figure 1.**
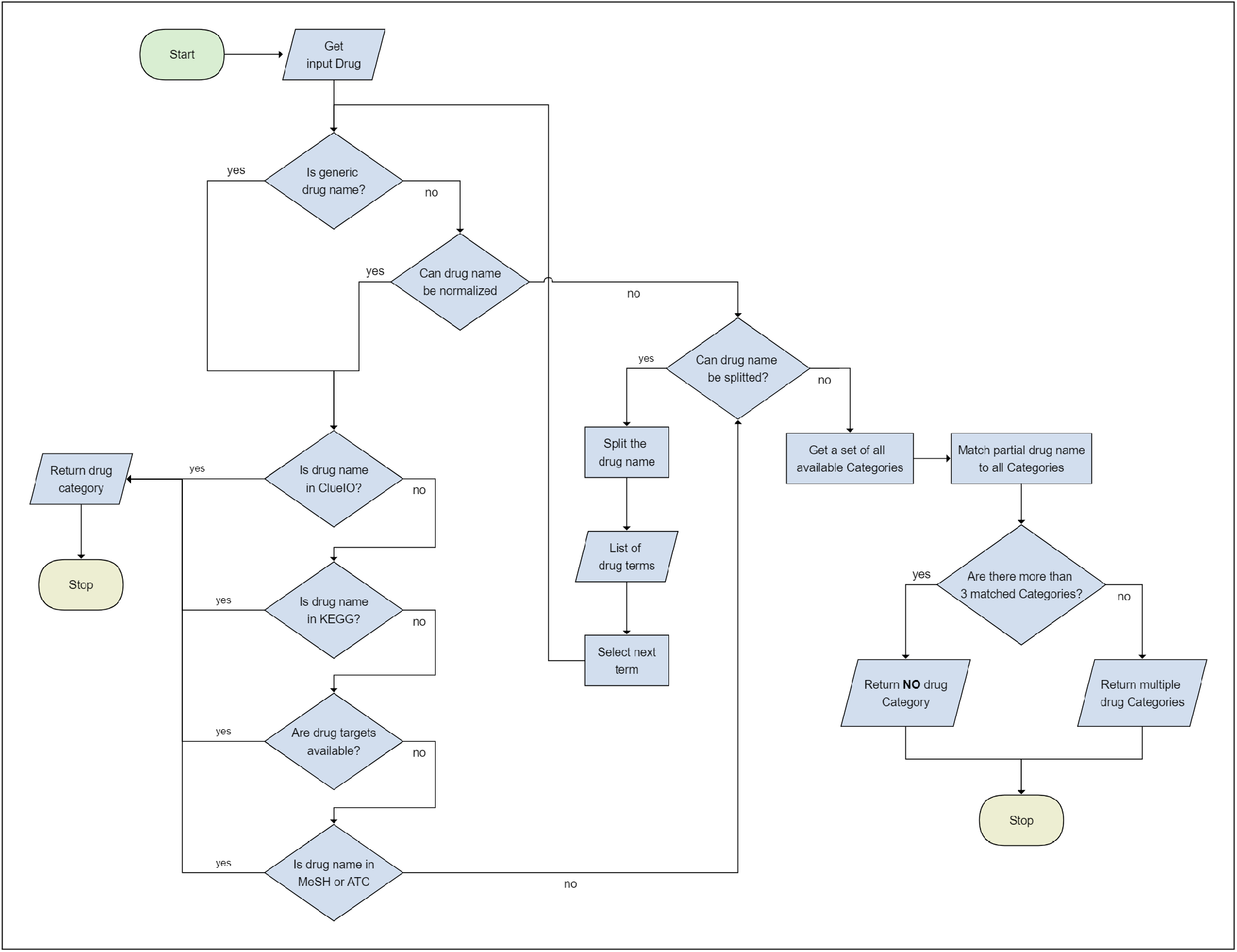
Automatic selection of general drug classes. The flowchart shows the steps DrugOn takes to process an entered drug to find the best match with a drug category.

The process initiates with the normalization of the input drug names to their generic equivalents. This includes the examination of the input name to determine whether it is a synonym or an identifier and the subsequent mapping of the name to the appropriate generic drug name. In the rare instance that this assignment is not feasible, the drug name is processed in a subsequent step for further processing if no initial categorization can be made.

Once normalization has been completed, a comparison of the drug name in question with other categorization databases is undertaken. The initial database queried is ClueIO, which was selected on the basis of its high degree of similarity to the manual classification. Should the drug in question be identified, the corresponding drug class is then retrieved directly. In the event that no classification is available from ClueIO, the search is expanded first to include KEGG Drug Class and KEGG Groups and continues with the suggested targets and mechanisms of action from DrugBank. The categories that correspond to the specified combination of target and mechanism (e.g., “PARP1” + “inhibition,” resulting in “PARP inhibitor” if it is available in other resources) are selected as the drug class. Ultimately, the accessibility of the drug name within the Medical Subject Headings (MeSH) or Anatomical Therapeutic Classification (ATC) databases is evaluated. Subsequently, these sources are consulted, as they provide a more expansive range of categories and a more comprehensive description of the drug.

In instances where the primary data sources are unable to provide precise information regarding a specific drug name, an alternative approach to the classification process is employed. This involves dividing the entered drug name into its individual substrings (e.g., “Selumetinib in BRAF-mutated tumors” becomes [“Selumetinib”, “in”, “BRAF”, “mutated”, “tumors”]). This method allows for the repetition of the preceding classification steps for each of the resulting parts.

In the absence of a precise match in the initial stages, the subsequent phase searches for partial matches across all available categories. The Levenstein distance is utilised to identify the most suitable matches, which are then filtered using a manually defined threshold. In the case of a high number of matches exceeding three, no category is assigned. In contrast, when three or fewer matches are identified, these are presented as probable drug categories.

### Software and Implementation

The ontology was created with the Python 3.9.7 programming language and the Owlready2 package (version 0.38), which is specifically designed for ontology-oriented programming. The software facilitates the creation of OWL ontologies in the form of classes and allows the inclusion of properties for the purpose of storing class-relevant information.

An application programming interface (API) has been developed to facilitate access to data from the ontology. The provided API utilizes the Flask package (version 2.1.2), a lightweight and robust web framework for Python, and incorporates a SwaggerUI documentation to enhance usability and accessibility. The documentation is available at https://mtb.bioinf.med.uni-goettingen.de/drugon/v1/doc/.

The front-end application has been implemented using Angular (version 16.1.4). Furthermore, the ontology-based answer (OBA) [24] server and OntoScope are incorporated for the graphical visualization of the ontology. It was designed with an intuitive interface to allow users to efficiently navigate and interact with the rich data stored in DrugOn. The application is accessible at the following URL: https://mtb.bioinf.med.uni-goettingen.de/drugon-web/ and the project’s source code is available at the following GitLab link: https://gitlab.gwdg.de/MedBioinf/mtb/drugon.

## Results

### Ontology Architecture

#### Ontology Classes

The structure of our recently developed DrugOn ontology hierarchy is based on three basic classes: Drug, Category, and Target (see Figure 2). These classes represent the fundamental elements of the DrugOn architecture, with each class fulfilling a distinct role in the organization of drug-related information. The entities of the domain are added as children to the ontology based on their respective basic class. All drugs are direct children of the base class drugs, other basic classes and the membership in drug classes and their targets are modeled with relationships to the children of the other basic classes.

**Figure 2.**
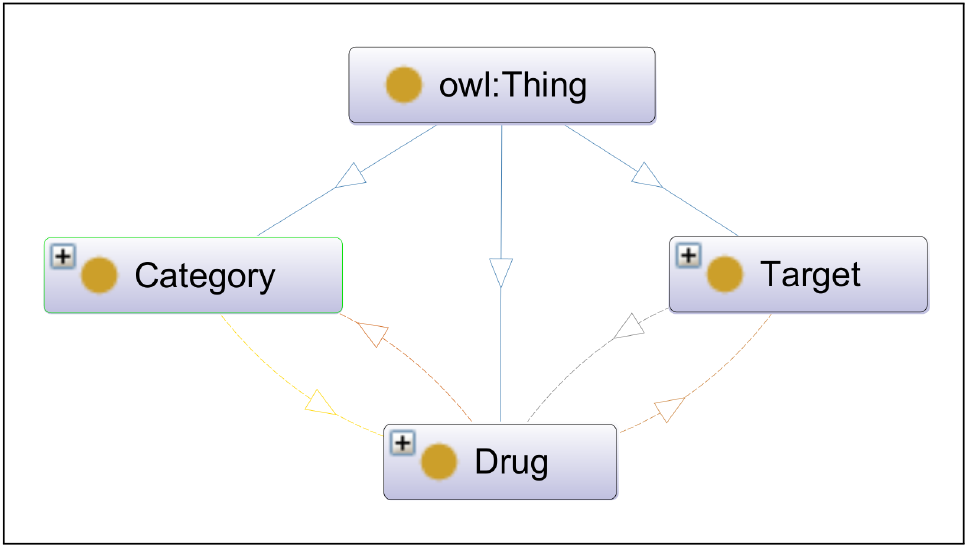
The principal architectural framework of the DrugOn ontology. The figure was exported by the ontology editor Protégé [25] and illustrates the basic classes, `Category, Target`, and `Drug`, along with their DrugOn relations. The drug classes are inherited from `Category` provided by the `is_a` relation. The relations to `Drug` are named after their originating sources and may be, for example, `has_atc, has_clue_io`, and so forth. The Genenames are inherited by the `Target` class. In relation to `Drug`, they indicate the action on these specific targets and are described as `is_inhibitor_for, is_antagonist_for`, e.t.c.

The drug terms identified on DrugBank are classified as subsets of the Drug class. A comprehensive representation of each drug is provided, including detailed information such as different identifiers (Chembl [26], ChEBI [27], KEGG [4], etc.) and synonyms. The `Target` basic class is concerned with the interaction between drugs and human proteins. It provides a link between the actions of drugs and biological entities. The MeSH terms, ATC codes, ClueIO annotations, [5, 6] and KEGG drug classifications [4], that describe a particular drug at various levels are children of the Category.

#### Relation Types

Relations are a fundamental component of ontologies, defining the functional connections between to classes named Domain and Range. Similar to classes specialization of relationships can be implemented by inheritance, also called sub-relations. They can also be defined as inverse, indicating the reverse relationship. In DrugOn the four basic inverse relation types in DrugOn are `targets/is_targeted, categorizes/is_categorizing`, each has subtype relations describing the specific relation and reflects the ‘Actions’-annotation in DrugBank. For example, the relation `[Drug] is_inhibitor_for [Target], [Drug] is_antagonist_for [Target] or [Drug] has_atc [Category]` can be seen in Figure 3.

**Figure 3.**
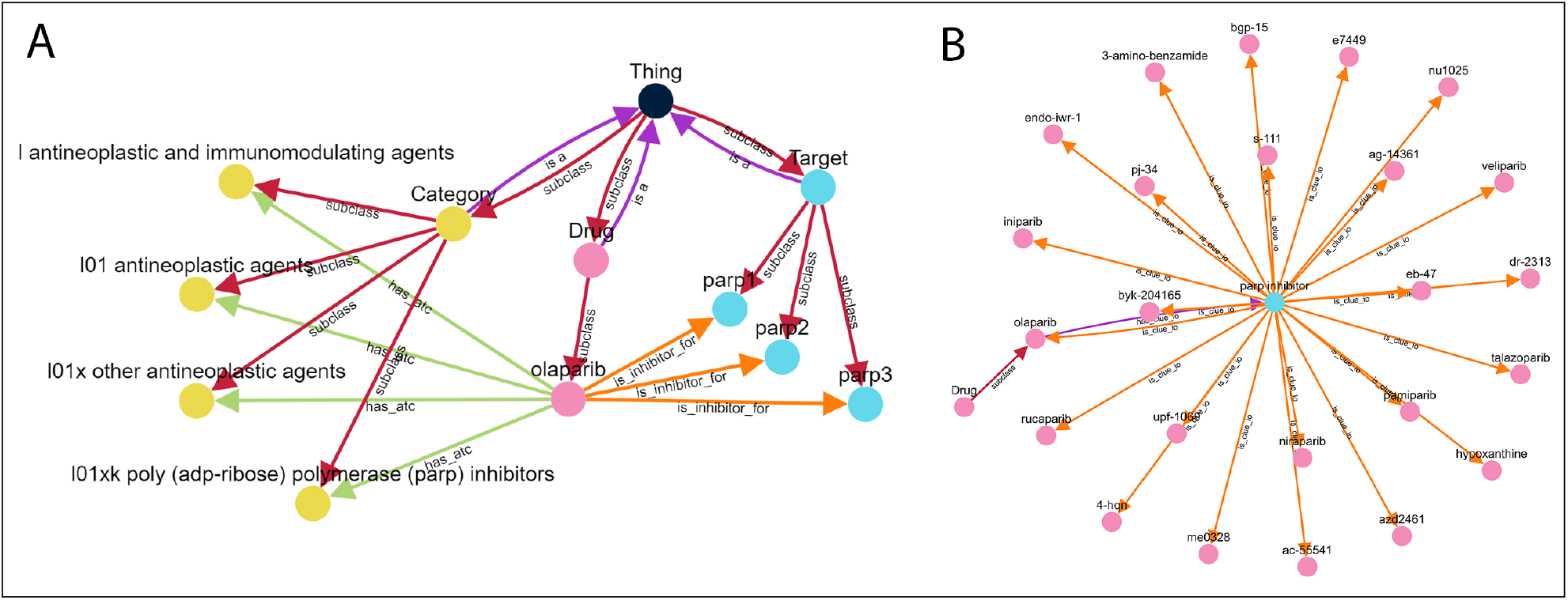
Ontoscope analysis in case of Olaparib. Ontoscope shows an example for the drug Olaparib. A) By searching for Olaparib, it is possible to expand the relationship ‘is_inhibitor_for’ (orange edges) to show all proteins that can be inhibited by the selected drug. It shows an inhibitory effect on the three targets PARP1, PARP2 and PARP3. By extending the relation ‘has_atc’ (green edges), Ontoscope shows all available categories from the ATC classification. This shows that Olaparib can be defined as an antineoplastic agent and a poly(ADP-ribose) polymerase (PARP) inhibitor. B) Search for alternative therapies with the same drug categories. In this case the selected database is ClueIO, which declares Olaparib as PARP Inhibitor -additionally there are 23 alternative drugs declared with the same category like Rucaparib and Niraparib.

The definition of these relationships provides a more detailed and accurate representation of a drug’s relation.

In conclusion, DrugOn’s architectural framework, with its hierarchical structure and comprehensive class relationships, provides a sophisticated platform for representing and analyzing drug information. Its capacity to encapsulate nuanced drug data, from biological targets to categorizations, renders it a valuable resource in precision medicine research.

### Implementation of the DrugOn Framework

The DrugOn framework consists of two main parts. The first one is constructing the ontology, based on Drugbank and enrich the classes with further information from ATC, MesH or ClueIO. Second the search algorithm to query the ontology. A significant challenge arises from the ability to input free text as therapies; this permits the use of non-generic drug names. For instance, we encounter in the therapy column ‘BRAF inhibitors In BRAF Mutant Tumors’, which is clearly understandable to humans, but it is challenging to automatically categorize this within an appropriate framework. DrugOn addresses this issue, by stepwise adjustment of the unmatched input (see Methods).

The DrugOn search can be accessed in different ways, depending on the desired scenario.

### Scenario 1 -General Drug Information

One of the primary feature of DrugOn is its web interface, which has been designed with the objective of facilitating user-friendly exploring of the ontology. The search for a drug offers access to all detailed information available in DrugOn. These include the automatic generated drug class, the drug’s mechanism of action, synonyms and identifiers, and all classifications made by other resources. This ease of use is of critical importance to researchers and clinicians who require rapid access to reliable and comprehensive drug information. Consequently, DrugOn represents a resource in both clinical and research settings, providing a readily accessible platform for obtaining essential drug information.

### Scenario 2 -Visual Ontology exploration

In addition to its search functionality, DrugOn offers an advanced interactive ontology viewer. This enables users to search for specific drugs, targets, and categories within the ontology, as well as to explore the complex relationships between these entities, thereby facilitating a deeper understanding of the relationships between various drugs, targets, and categories.

One of the key capabilities of this feature is the ability to expand queries to include related nodes. By expanding the ontology by a selected relation type the user can walk through the ontology according to a specific question. This advanced query functionality is particularly valuable for understanding complex drug-target combinations and the nuances of categorization within the ontology.

For researchers and clinicians, this provides a comprehensive understanding of the interactions between different drugs and their targets, as well as the classification of drugs within the ontology. This enables a deeper understanding of the mechanisms of action, potential adverse effects, and therapeutic options of various pharmaceutical agents. The capabilities of DrugOn significantly enhance its utility as a tool for both clinical and research applications.

### Scenario 3 -API and OWL-File for automatic data analysis

In order to facilitate large-scale data analysis, Drug-On includes an API that provides automated access to its classification functions as in the other described use cases. This API is crucial for integrating Drug-On’s functionalities into broader data processing workflows. Researchers and clinicians can utilize the API to standardize drug names, perform synonym or identifier queries, and extract information about drug classes, targets, or other relevant attributes.

The API supports various endpoints that streamline interactions with the ontology, enabling efficient data handling and analysis. For example, users can query the API to retrieve all drugs associated with a particular mechanism of action or to assign drugs according to their corresponding class. This automation reduces the manual effort required for data processing and ensures consistency across large datasets.

DrugOn is implemented using the Web Ontology Language (OWL), a formal specification that facilitates comprehensive and machine-readable representations of knowledge. The OWL ontology file serves as the fourth access path to DrugOn. It can be used for other tools or in environments with restricted internet access.

In summary, DrugOn offers a comprehensive range of features, including the provision of detailed drug information through a user-friendly web interface and the possibility of undertaking detailed analysis through its API or locally usable OWL ontology file. Together, these features enhance DrugOn’s applicability in a variety of research and clinical scenarios, making it a valuable tool in the field of drug analysis.

### Comparison: DrugOn vs manual annotation

DrugOn was tested on 336 drugs applied in translational lymphoma research, spanning all available therapy types in CIViC. A comparison of the automatic classification with a manual, database-derived annotation reveals a high degree of correspondence and provides more detailed supplementary information. 54 drugs could not be assigned a classification because no information was available in the underlying data resources or inconsistent manual text entries in the input data. These 17 out of 54 entries failed due to descriptions that were difficult to interpret by machine. For example, descriptions such as ‘corticocorticosteroids In Early Setting’ or ‘MEK inhibitors In BRAF Mutant Tumors’ appear in CIViC, which are clearly understandable for humans, but present a challenge for machine interpretation. In the remaining 282 drug descriptions, the automatic classification returned correct descriptions or matched the manual descriptions. An excerpt of the classifications can be found in Table 1. The complete table is accessible via the supplementary data and contains the name of each drug, a manual annotation, DrugOn’s classifications, and the drug’s target proteins, including the mechanism of action.

DrugOn’s robust performance in accurately classifying a diverse array of CIViC therapies highlights its capabilities. This is not limited to the current version of CIViC but should be also continued with newer versions of the used primary resource, and for other data catalogues, similar to CIViC. However, this functionality represents merely one facet of DrugOn’s utility. It offers a multitude of additional features that address a spectrum of use cases, thereby further enhancing its pivotal role in drug information management.

## Discussion

In the evolving landscape of precision medicine and drug research, we developed DrugOn as a crucial advancement. It demonstrates the power of integrating different databases to create a multidimensional ontology that combines the best features of each resource into a single repository. This innovative approach not only simplifies drug classification, but also improves the accuracy and applicability of drug data in various biomedical projects.

The graph based nature of ontologies allows to select the appropriate relation type according to the use case like molecular tumour boards or research projects. Similar DrugOn provides different access options for the use cases such as web based research for manual preparation of the MTB or the API for bioinformatic supported projects. The automatic construction of DrugOn facilitates an up-to-date integration of the included resources. By providing easy access to data on drug classes, targets and similar drugs, DrugOn significantly simplifies the decisionmaking process for clinicians.

In addition, DrugOn’s unbiased methodology ensures that the information provided is not skewed by individual experience or biases towards the desired effect, thereby increasing the objectivity and reliability of drug classifications. The use of DrugOn in projects such as the mapping of drug classes in lymphoma data projects and its integration into Onkopus, a framework for cancer variant interpretation in the scope of a MTB (https://mtb.bioinf.med.uni-goettingen.de/onkopus/), exemplifies its versatility and importance. In these applications, DrugOn’s ability to automatically categorize treatments and present comparable options allows for a more nuanced and informed approach to support therapeutic strategies.

Existing drug hierarchies such as the ATC (https://atcddd.fhi.no/) and DRON [19] cover different ranges of hierarchies. For example, ATC is well suited for basic levels, while DRON focuses on drug application and pharmaceutical details. In DrugOn, we integrate multiple data sources to create a comprehensive drug information retrieval tool. Compared to similar approaches, DrugOn has both advantages and limitations. One of its unique features is an ontology-based data source that supports multiple scenarios and allows visual inspection of therapeutic options. Other resources, such as the NCI-Thesaurus (https://ncit.nci.nih.gov/ncitbrowser/) and DRON, also use multidimensional hierarchies. However, DrugOn emphasises a user-friendly graphical ontology browser designed for interactive exploration of research questions.

DrugOn also addresses the needs of different research areas by offering different access options. In translational research projects, users can start with the web interface in the early stages of research and later switch to the API for large-scale data processing. Other drug classification systems, such as ATC and DrugBank [7], provide broad coverage of drugs across all areas of medicine. However, DrugOn’s primary focus is on precision oncology and it includes all DrugBank drugs in its database. In particular, DrugOn offers advanced features that highlight molecular interactions of chemotherapeutic agents.

The automatic generation of DrugOn’s ontology has highlighted the need for high quality input data. For example, when evaluating the CIViC [23] data mappings, a significant number of drugs could not be assigned because the free text annotations did not contain the correct drug names. Another class of unmatched drugs are those for which there is insufficient data in DrugBank. Future versions of DrugBank are expected to address these gaps. However, the field of oncology is rapidly evolving and new drugs are constantly being developed [13].

DrugOn is a detailed and easy-to-use platform that significantly improves the ability to classify and use drug data and explore alternative treatment options. In short, DrugOn improves the way we use drug information in precision medicine. DrugOn’s success in projects such as clinical decision support shows its potential to contribute to healthcare by making information more accessible and reliable.

## Data Availability

All data produced are available online at: https://gitlab.gwdg.de/MedBioinf/mtb/drugon

## Author Contributions

Idea and Design: KK, JD, RK, TB. Backend implementation: KK, VG, MH. Frontend implementation: KK. Data annotation: RK. Data validation: KK. Paper writing: KK JD; All authors approved the manuscript in the submitted version and take responsibility for the scientific integrity of the work.

## Acknowledgement

We thank the Göttingen Promotionskolleg für Medizinstudierende for the support of Myrine Holm.

